# Virtual mindfulness interventions to promote well-being in young adults: A mixed-methods systematic review

**DOI:** 10.1101/2021.04.24.21256035

**Authors:** Joy Xu, Helen Jo, Leena Noorbhai, Ami Patel, Amy Li

## Abstract

**Background:** With the onset of the COVID-19 pandemic, students have experienced drastic changes in their academic and social lives with ensuing consequences towards their physical and mental well-being. The purpose of this systematic review is to identify virtual mindfulness-based interventions for the well-being of young adults aged 15 to 40 years in developed countries and examine the efficacy of these techniques/exercises.

**Methods:** This mixed-methods systematic review follows the Preferred Reporting Items for Systematic Review and Meta-Analyses (PRISMA) guidelines with a registered PROSPERO protocol. With a convergent integrated synthesis approach, IEEE Xplore, PsychInfo, Web of Science and OVID were searched with a predetermined criteria and search strategy employing booleans and filters for peer-reviewed and grey literature. Data screening and extraction were independently performed by two authors, with a third author settling disagreements after reconciliation. Study quality of selected articles was assessed with two independent authors using the Mixed Methods Appraisal Tool (MMAT). Studies were analyzed qualitatively (precluding meta and statistical analysis) due to the heterogeneous study results from diverse study designs in present literature.

**Results:** Common mindfulness-based interventions used in the appraised studies included practicing basic mindfulness, Mindfulness-Based Stress Reduction (MBSR) programs, Mindfulness-Based Cognitive Therapy programs (MBCT) and the Learning 2 BREATHE (L2B) program.

**Conclusion:** Studies implementing mindfulness interventions demonstrated an overall improvement in well-being. Modified versions of these interventions can be implemented in a virtual context, so young adults can improve their well-being through an accessible format.

## INTRODUCTION

Well-being is a subjective state without a universal definition due to its diverse manifestation in different individuals. According to the Centers for Disease Control, well-being can be defined as having a positive outlook on life and feeling content [1]. Beyond the mental aspect, well-being encompasses a holistic approach to health by integrating both physically and mentally positive aspects throughout daily activities [1]. Thereby, well-being serves as an important factor in maintaining the quality of life for all individuals.

Within academia, well-being is a particularly vulnerable area as students transition through varying stressors, including changes in lifestyle, academic stress, and new responsibilities. The transition from adolescence to adulthood presents additional risks for poor well-being with potential consequences affecting one’s academic and personal life [2]. In 2019, the American College Health Association found 36.5% of US college students reporting stress as a major factor negatively affecting student’s academic performance [3]. Amidst the COVID-19 pandemic, students face additional challenges; 71% of students from American public universities report a direct increase to stress levels due to the pandemic, which also contributes to more frequent depressive thoughts and higher levels of anxiety [2]. Thus, there is a clear need for improving student well-being through personalized interventions. For students, these interventions may assist in managing stress levels, reducing anxiety, and improving cognitive awareness (improves learning skills and productivity).

Mindfulness is a significant aspect of well-being. Mindfulness involves bringing awareness to present-moment experiences like thoughts, body sensations, and the environment (without judgement). Mindful awareness contrasts with the default mode of everyday life, where the state of attention for many individuals is inattention as the mind may wander or operate mindlessly. However, awareness of the present requires one to consider internal experiences including personal thoughts, feelings and sensations, which may elicit attention towards potentially negative emotions that certain individuals tend to suppress. For many individuals, the ability to confront and process negative thoughts and emotions is a difficult process. These aversive effects are illustrated by 11 laboratory studies, where healthy adult subjects preferred monotonous tasks or mild electrical shocks rather than being left alone in their own thoughts [2]. However, mindfulness provides an opportunity for individuals to acknowledge these aspects and progressively improve habits or lifestyle choices. Bringing awareness to negative thoughts as they arise prevents them from spiralling out of control into further negative thought patterns. By implementing personalized interventions, the transition towards practicing mindfulness can be eased to reduce negative responses to one’s internal and external experiences.

Mindfulness provides long-term management of well-being as individuals become aware of the root of negative emotions and resolve these issues rather than let them worsen in the future. With greater awareness of the present, individuals may confront and accept feelings of stress, anxiety, fear, hate, sadness with greater ease. Unhealthy ways of coping, suppression of stress, mental breakdowns, burn out, and physiological issues from chronic stressors can result from maladaptively addressing these negative emotions [2]. The significance of mindfulness interventions are explored within research, but a gap in the present literature remains within virtual integrations of these interventions specifically targeted towards young adults.

Therefore, this systematic review assesses effective strategies to improve well-being within an academic context for young adults from ages 15 to 40 years old. By identifying, appraising and integrating present literature, mindfulness-based interventions can be evaluated for most desirable outcomes. This mixed methods systematic review aims to answer the following research question: What mindfulness-based interventions can be integrated into a mobile application to promote well-being of individuals (students) between the ages of 15 to 40 years old? These findings may be integrated into the *Felicity App*, a virtual application targeting students in developed countries to provide productivity enhancement techniques through a mobile and web application. Productivity incorporates several facets, where well-being serves as an important aspect to improve productivity and proactively prevent demotivation and burnout by managing stress levels.

## METHODS

A systematic review was conducted to assess mindfulness interventions in the context of well-being for students to improve productivity, motivation, and overall mental health. The current review was conducted and reported in accordance with the Preferred Reporting Items for Systematic Reviews and Meta-Analyses (PRISMA) statement [67] and the protocol is registered in PROSPERO, an international database of prospectively registered systematic reviews in health and social care.

### Study Design and Setting

Virtual mindfulness interventions that can improve students’ well-being were examined through this systematic review. The virtual element of these interventions may allow for multi-media incorporation, such as within a mobile application to help students improve their emotional and academic experience. Apps allow for interactive participation, accessibility and flexibility of options. Statistics Canada has determined that many people 15 years of age and older own a smartphone and 45.4% of those individuals check it every 30 minutes [4]. Thus, a smartphone app is accessible and virtual interventions through mobile apps can be seamlessly integrated into the lifestyles of this age demographic. The target audience for this application are young adults between 15 and 40 years of age in developed countries located in North America, Asia, Australia and Europe.

A mixed method study design was used to ensure qualitative and quantitative elements of the mindfulness-based intervention strategies are included in the systematic review. Quantitative data provides measurable information on how mindfulness-based interventions improve cognition and overall well-being for students. Qualitative data allowed for identification of patterns observed with mindfulness-based interventions and the perspectives associated with different intervention methods. Both forms of data are important to ensure the most effective mindfulness-based interventions are integrated and that optimal well-being strategies can be used by users. Due to the diverse study designs, subsequent study findings have shown significant heterogeneity and statistical analysis is precluded by qualitative analysis.

### Search Strategies & Criteria

IEEE Xplore, PsychInfo, Web of Science, ProQuest and OVID were searched for articles published between January 1980 to January 2021, using keywords related to mindfulness, productivity, wellness, and cognition in students. A broad range of synonyms were used to ensure all relevant studies were included in the search, with accompanying filters and booleans (refer Appendix 1 for the full search strategy and Appendix 2 for the grey literature search strategy).

For the implementation of grey literature, a search for articles was conducted in various databases and websites, specifically the National Center for Biotechnology Information (NCBI) and the Web of Science. The snowballing method was used to search Google Scholar to identify additional sources from references cited in conference proceedings and web pages.

The search strategy was created by using keywords with different booleans and truncations. The specific words “mindfulness and student*, learn*, burnout or stress or motivat*, productiv*, cognit*, atten*, creativ*, virtual, intrinsic were used for extraction. Other specific words such as teacher or professor or kindergarten or children or ADHD or GAD or TBI or injury aided in excluding material through the “not” function. This comprehensive set of sources were processed with more advanced screening to attain the most relevant literature. Specific filters were added to databases to screen full-texts when possible.

### Eligibility Criteria

The predetermined criteria for inclusion includes psychology studies published in English between 1980 to January 2021. Eligible study designs included randomized controlled trials (RCTs), meta-analyses, systematic reviews, and grey literature. To be included, studies must mention the impact of mindfulness interventions on any of the following: motivation, burnout, stress, learning, cognition, and productivity related topics. These interventions must be virtually transferable and excluded individual therapy. The study participants were young adults between ages 15 to 40 of all genders, and residing in North America, Europe, Asia, or Australia. Studies with interventions targeted to individuals with a physical illness, mental illness, or learning disability were excluded. Any articles that were not available online and were published by organizations with conflicts of interest were also excluded from this systematic review (refer to Appendix 4 for a list of excluded studies).

### Data Collection and Abstraction

A predetermined, piloted screening tool was developed using the inclusion and exclusion criteria. The screening tool ensured that only relevant data would be collected from abstracts and full text articles. The pilot trial of the screening tool prompted necessary revisions to improve precision. Modifications to the tool were prompted by discussions during reconciliation.

A total of 915 articles were initially searched, with findings consisting of 71 grey literature and 844 peer reviewed articles. The database search results were combined, and duplicate articles (n=20) were removed manually using Endnote (version 8) prior to the screening phase. Within study selection, titles and abstracts of 895 articles were screened by two independent authors on January 31, 2021. Any disagreements between the two independent authors were reconciled and disagreements were resolved with a third author. Prior to full-text screening, 3 grey literature and 116 peer-reviewed articles proceeded towards full-text screening with two independent authors, reconciliation, and a third author for resolving disagreements. After full text screening of the initial papers, a total of 22 studies remained.

Upon initial study selection, reference lists of included articles were hand searched and screened for potential inclusion. During the first round of hand search screening, 1302 peer reviewed articles went through abstract screening by two independent authors using the piloted screening tool to produce 57 papers. Full texts were screened independently by the two authors, then reconciled to resolve any disagreements with a third author for resolution. In total, 17 studies from hand searches were included. Data extraction was performed in duplicate with two independent authors, with study characteristics including author, title, study duration, study population, location, intervention methods and results. In total, 39 studies were included in this systematic review.

### Quality Assessment

The Mixed Methods Appraisal Tool (MMAT) was used to assess the quality of the articles included in the study. The MMAT is a critical appraisal tool used to assess qualitative, quantitative, and mixed methods studies. The MMAT provides an effective measure to appraise a wide variety of empirical studies. The tool has been tested for reliability and content validity, however, the literature on the quality of the MMAT lacks consensus. Articles are organized into five categories: (1) qualitative research, (2) randomized controlled trials, (3) non-randomized trials, (4) quantitative descriptive studies, and (5) mixed methods studies. Upon determining study category, quality ratings are provided according to subsequent criteria.

Two independent reviewers appraised 39 articles using the MMAT. After individually assessing the articles, reviewers reconciled ratings and a third author resolved conflicts through discussion. Duplicate studies were removed after the review and a total of 32 quality appraised studies remained.

### Data Extraction

Data extraction was completed by two independent reviewers and reconciled, with a third author for potential disagreements. A table was synthesized for the final study characteristics (refer to Appendix 3). The table consisted of the following information: first author, title, country, study design, duration, participants, type data, outcome, and quality/design score. The quality/design score was obtained through the process of quality assessment using the MMAT.

## RESULTS

### Study Characteristics

The study selection and screening process is outlined in Fig. 1. A total of 32 studies are included in this systematic review. Of all the articles, 3 were qualitative studies, 5 were quantitative non randomized studies, 4 were quantitative descriptive studies, and 3 were mixed methods studies. The remaining 17 studies were quantitative randomized controlled trials. Of the 32 articles, Netherlands, Singapore, Japan and Australia contributed one study per country. 2 studies were from Taiwan and the remaining studies were from the USA. All included studies passed the initial screening criteria and was rated based on the MMAT. Among the 32 articles that were quality appraised 12 were rated an overall score of five, 16 were rated four, and 4 were rated three. None of the included studies were rated two or one stars.

**Fig. 1.**
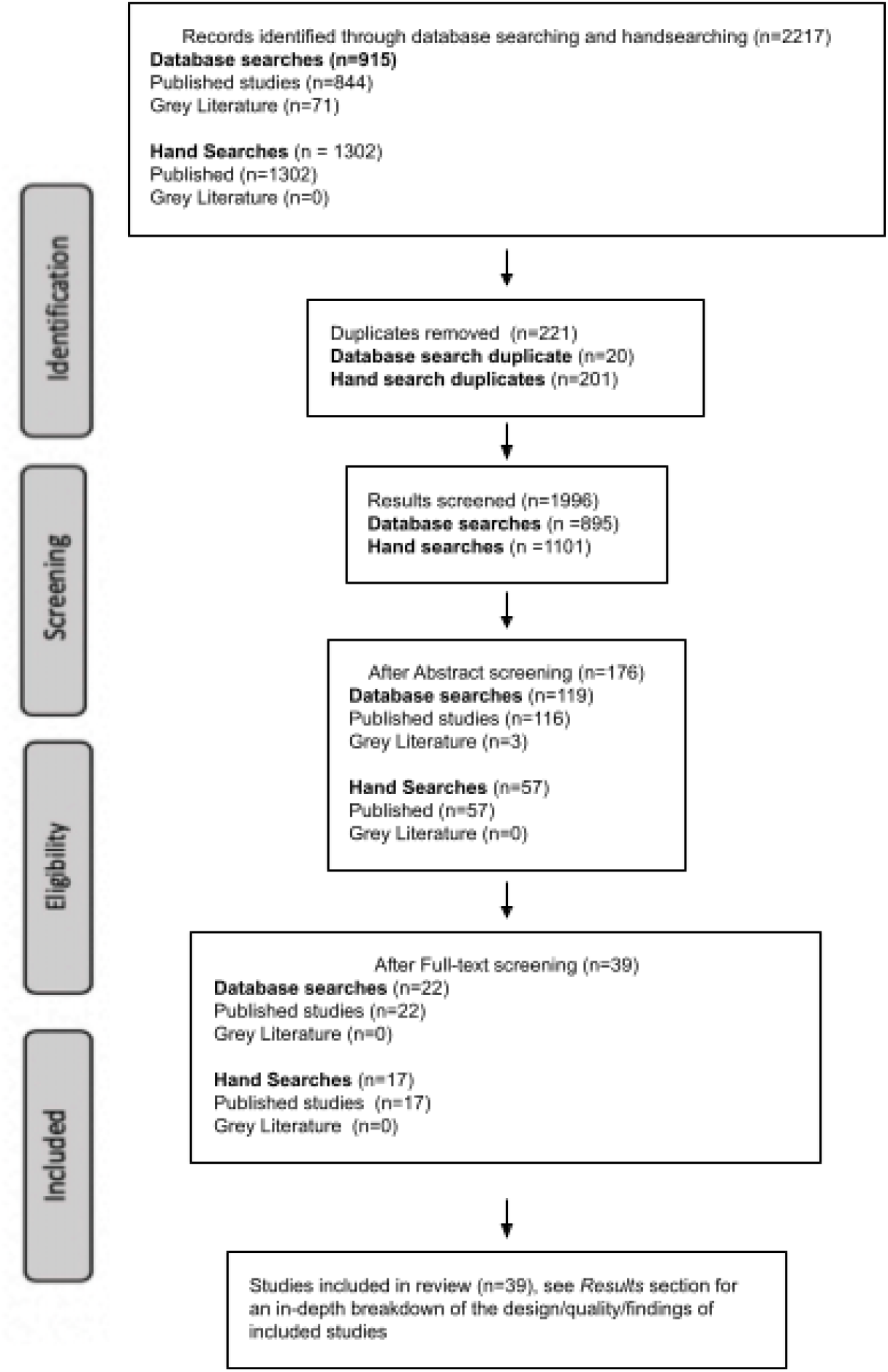
Structural diagram of the screening and study selection processes completed

### Study Outcomes

Data and results were extracted from the literature and synthesized into a study characteristics table (refer to Appendix 3). The main outcomes that met the inclusion and exclusion criteria and contributed to improved well-being were compared. The analysis of mindfulness interventions and their effectiveness resulted in the identification of five themes.

1. *Basic mindfulness* refers to an intervention involving focused attention to a sensory experience such as rhythmic breathing and attention monitoring.
2. *MBSR* refers to an 8-week Mindfulness-Based Stress Reduction program encompassing techniques for guided mindfulness using meditation, and a focus on opening up to a moment-by-moment experience.
3. *L2B* refers to the Learn 2 Breath program dedicated to the acronym *BREATHE*. The core themes established include: having a body awareness, understanding one’s thoughts, working with one’s feelings, the integration of thoughts, feelings and sensations, decreasing harmful self-judging thoughts, and making a conscious effort into integrating mindfulness practises in daily life.
4. *MBCT* refers to mindfulness-based cognitive therapy which focuses on acceptance and awareness of thoughts and emotions.
5. *Unique Methods* included different techniques that did not fit into established programs and were used for assessing positive emotions, feelings of meaning, and purpose.

### Intervention Targets

The five qualitative mindfulness themes that contributed to improving well-being were found to be interrelated and often had similar targets and outcomes.

#### Overall Mental Health Symptoms

Strong correlations were determined between the implementation of mindfulness interventions and overall mental health. Students that practiced mindfulness reported lower levels of stress, depression, and sleep issues, leading to an overall increase in the well-being of young adults [5]. Mindfulness interventions also decreased the frequency of experiencing negative emotions and enhanced attention to personal emotions [6]. Heightened awareness resulted in better regulation of their emotions so that students could experience greater equanimity and less reactivity [6]. In addition, students practicing mindfulness reported significantly less burnout compared to the control group [7].

#### Anxiety

Anxiety is a common symptom observed in young adults experiencing constantly changing environments, particularly in individuals learning how to adapt to varying work flows and lifestyles [8]. The effect of different mindfulness-based interventions (MBI) were assessed to observe its effect on improving anxiety in college students. A study by Bamber et al. [9] concluded that MBI with a focus on relationships/loving-kindness and insightful meditations were shown to have no significant effect on anxiety. However, students who participated in brief focused breathing exercises reported increased performances in difficult arithmetic tests, due to a reduction in anxiety after the exercises [10]. When students participated in MBI, a reduction of anxiety symptoms was reported [11, 12]. These studies indicate that MBI with a focus on breathing can reduce anxiety levels in college students.

#### Academic Performance

MBI has also been shown to influence academic performance in students. Bellinger et al. [13] determined an indirect benefit of mindfulness in improving math performance by reducing anxiety. Mindfulness interventions before quizzes improved performance, but short five minutes bi-weekly sessions of mindfulness had no significant effect on students’ exam scores [14]. These findings suggest that brief mindfulness can be beneficial for enhancing knowledge retention of lecture content in the short term. Lin & Mai [15] also found significant short-term improvements in academic performance. In the long-term, however, several studies found no significant improvements in academic performance after mindfulness interventions [12, 16]. In contrast, Cavanagh et al. [17] found that mindfulness interventions resulted in better final exam performance in conjunction with cognitive reappraisal (which involves mentally reframing views of boredom, frustration, and anxiety to change their meaning and the emotions they arouse). Though mindfulness may not have directly impacted exam performance, mindfulness could increase student awareness of personal feelings (such as boredom) for recognition and proactively apply cognitive reappraisal techniques to reframe them.

#### Cognition

Mindfulness interventions played a role in enhancing metacognitive ability. The implementation of meditation exercises were shown to enhance introspective accuracy including visceral sensations, affective states, and ongoing performance on tasks. Meditation exercises focused on breathing enhanced metacognitive ability for memory, but there were no significant improvements in perceptual decisions [18]. Although many papers reported that mindfulness led to more positive emotional experiences, interventions did not produce significantly higher levels of emotional intelligence [7].

In addition, interventions reduce mind wandering on tasks requiring sustained attention which may be beneficial for college students, as most schoolwork requires sustained attention [19]. Students also report being more “on-task” after mindfulness training [19]. Yamada and Victor [16] found that 81% of students self-reported positive effects of mindful awareness practices on their learning, indicating that students enjoy this practice.

In terms of memory, a short-term 15-minute mindfulness practice had no impact on working memory tasks [21]. Working memory refers to the ability to temporarily store and use the information to solve problems. A week-long mindfulness intervention did not increase working memory or decrease mind wandering, yet it prevented stress-related working memory impairments, which suggests short-term mindfulness has indirect benefits to working memory [22]. However, a more extended approach where focused attention meditation was practiced for two weeks showed significant improvements in memory [18].

## DISCUSSION

This systematic review provides an examination of the current literature regarding the impact of mindfulness interventions to improve well-being along with academic success. Generally, practicing mindfulness improved mental health and had positive psychological results, but the outcomes for academic performance were mixed.

Five different intervention strategies will be discussed in depth for potential implementation. Considering the COVID-19 worldwide pandemic, it is important that these strategies can be applied successfully in virtual settings as well.

### Common Mindfulness Interventions

#### Basic Mindfulness

Many studies included in this review incorporated generic mindfulness techniques rather than following specific programs. Common exercises included in sessions are rhythmic breathing, meditation, attention monitoring, and acceptance. Baird et al. [18] identified basic mindfulness as meditation exercises requiring focused attention to some aspect of sensory experience. Wei Lin et al. [15] introduced basic sitting meditation to students to stabilize the mind and rhythmic breathing exercises to increase focus and self awareness. Rhythmic breathing involves focusing the attention on one’s nostrils, and the act of breathing [15]. Results from this study showed that those who completed these basic MM were able to sustain their focus, improving learning.

Mindfulness questionnaires also indicated that students felt an increased amount of mindfulness after completing the meditations, compared to the control group [15]. Other forms of basic mindfulness focus on attention monitoring and acceptance. Attention monitoring trains students to focus on breathing and somatic sensations, thoughts and emotions, and meta-awareness of cognitive, emotional and physical events [19]. Acceptance trains students to have non-judgemental attitudes towards these thoughts and sensations [19]. Many other studies were conducted in which students were trained in these MM practices as an anchor for attention, so that they could sustain their attention on sensations and emotions without being distracted [18]. Generally, implementations of these techniques enhanced students’ ability to sustain attention which improved short-term academic and task performance [6, 12, 23, 24, 25].

#### Mindfulness-Based Stress Reduction(MBSR)

A common mindfulness intervention that is effective for improving college student well-being is the Mindfulness-Based Stress Reduction (MBSR) program lasting 8 weeks [7]. MBSR is a form of guided meditation to improve mindfulness. Jain et al. [26] examined the effect of the MBSR program on full-time medical students, nursing graduate students, and undergraduate students enrolled in a premedical program. The study compared mindfulness meditation (MM) and somatic relaxation (SR) given in the MBSR format focusing on the improvement of students’ well-being. The MM intervention consisted of guided body scan meditation, where one gradually focuses their attention to each part of the body, sitting meditation, Hatha yoga (which consists of stretching the body through gentle movements), walking meditation, and loving-kindness meditation, where one expresses feelings of love and kindness to oneself and others [26]. The SR intervention consisted of muscle relaxation and breathing exercises. The results indicate that MM interventions given in the MBSR format enhance positive mindsets and reduce ruminative and distractive thoughts associated with depressed moods, increasing cognitive ability [26]. Another study was conducted on medical students using the MM interventions stated in the previous study [5]. Medical students were given training in all of the interventions over 8 weeks. The MM interventions given over the MBSR program in this study were shown to improve psychological well-being, reduce state and trait anxiety, and increase empathy [5].

A derivative of MBSR was used to assess the effect of mindfulness practices on student learning in a course. The study consisted of a 10-minute sitting meditation routine led by an instructor at the beginning of each class [16]. Students in the intervention group reported a better cognitive experience by participating in the 10-minute mindfulness activity before class [16]. However, the final exam scores showed no significant improvement for the course between the intervention group and the control group without training [16]. Studies were also conducted comparing MBSR and E. Easwaran’s 1978/1991 eight-point program (EPP) on improving mindfulness in college students. No significant differences were found between MBSR and EPP, and both were found to reduce stress [26, 27]. An 8-week long MBSR program with 7 weekly two-hour classes was set up for students [28]. The group which participated in the MBSR program showed significant improvements on the Mindfulness Attention and Awareness scale, and improvements with working memory capacity and visual threshold [16]. Sankoh [7] created a modified 8-week MBSR course for medical and premedical students with reduced class times to match the students schedules. Students were also encouraged to use the Headspace mobile app to practice mindfulness during their own time [7]. Little significance was found in mindfulness scores between students who participated in the modified MBSR compared to those who did not [7]. Those who did the course reported feeling smaller impacts of burn out, compared to those who did not do the course [7]. Articles in this review show strong evidence that MBSR programs can be an effective method for students to increase mindfulness and reduce stress [7]. They can also be modified to fit the lifestyles of students, and still have a significant improvement on well-being.

#### Mindfulness-Based Cognitive Therapy (MBCT)

Mindfulness meditation can be taught in the form of Mindfulness-based cognitive therapy (MBCT). The goal of Mindfulness meditation (MM) was to promote awareness and acceptance of thoughts and emotions rather than the suppression of them [22]. Participants were taught to focus on breathing and take note of any mind wandering that occurs, promoting acceptance, rather than judge oneself for it [22]. Focusing the mind on simple breathing was encouraged [22]. Results of this study indicated that MM did not improve mind wandering, or working memory, but did have a strong significance in improving mindfulness in students [22].

#### Learning To BREATHE (L2B)

A popular intervention revolved around specific focused-attention meditation and breathing called Learning to BREATHE (L2B) [17]. In these techniques, students were told to choose a specific task (such as breathing), recognize when the brain becomes distracted from the task, be able to shift attention back to breathing, and have strong cognitive understanding of distractors [14]. Core practises included a body scan, maintaining mindfulness of thoughts, emotions and movements [29]. These breathing exercises were successful when performed right before small quizzes, but not in long-term effects on scores [29]. Additionally, the goals for the program involved gradually building up inner strength and empowerment through the commonly used structure of 8 sessions [29]. The program hoped to enhance the students’ skills in regulating emotions, improve management of stressful situations, and improve learning processes [29]. As a result, this program was successful for stress reduction and management, self-regulation, and improvements towards a healthier lifestyle (e.g. more exercise and less alcohol consumption) [30]. Finally, participants in the program demonstrated high gains in emotion regulation above all [29]. Overall, L2B is an effective method for long term emotional/stress regulation.

### Unique Methods

Some papers presented unique mindfulness interventions that were applicable to their specific research aims without using pre-existing programs. Instead of using general mindfulness techniques, studies used cognitive reappraisal interventions, positive psychology interventions (PPIs), and somatic psychoeducation [17, 31, 32]. Through cognitive reappraisal interventions, there was a focus on normalizing emotions in a college classroom setting [17]. Mindful ways of approaching emotional experiences were introduced, rather than techniques for reappraisal [14]. PPIs were used for students taking a positive psychology course [31]. One study researched the effects of a somatic psychoeducation experimental course on well-being by organizing activities designed to increase somatic awareness and trust between other people [32].

### Implementations

One way that mindfulness interventions can be introduced into the *Felicity App* is through breathing reminders. Users can choose to receive reminders to breathe and focus their attention on body sensations and present emotions. Notifications to ‘remember to breathe’ could appear hourly or whenever the user decides upon routinely. The pop up reminder could also include a link to more detailed meditation practices or an image with a quick exercise. One limitation is that pop up notifications are easily dismissible.

Another feature that can be incorporated is modified versions of pre-existing mindfulness programs, like MBSR (8 weeks) and L2B (6 weeks). The programs introduce different mindfulness related topics weekly so the same topics could be presented in the virtual application as simplified exercises. Guidance on activities could be given through an audio recording or through written instructions where students will be completing tasks in an individual setting. Activities that could potentially be included are body scan meditation, hatha yoga, walking meditation, and loving-kindness meditation. Each activity is useful for targeting symptoms influencing the well-being of students.

### Strengths and Limitations

Literature with varying study designs were included for this systematic review, resulting in high heterogeneity precluding statistical analysis. Incorporating qualitative, quantitative, and mixed method studies enabled a qualitative analysis of present data. Self reports and surveys were analyzed to gain more insight into how mindfulness interventions impact individual experiences and overall well-being.

This systematic review included articles with participants from North America, Europe, Asia, or Australia but excluded other countries, which may lead to potential bias in data. Of the 26 studies from North America, all studies were conducted in the US. 4 studies were conducted in Eastern and Southern countries of Asia, specifically Taiwan, Singapore and Japan. Only 1 study was conducted in Europe. Studies only from one country may be a poor representation of the region as a whole. In addition, only studies written in the English language were included in this systematic review which can narrow the scope of literature findings. Due to the heterogeneity of study design and findings, further research (including additional quality assessment and statistical analysis) is required to determine the efficacy of virtual mindfulness interventions within well-being.

## CONCLUSION

In this systematic review, the effect of different mindfulness-based interventions on the well-being of college students was studied. The symptoms to be targeted by mindfulness-based interventions were investigated, as well as the procedures and outcomes of the interventions. Specific criteria were used to include articles relevant to the application of implementing mindfulness-based interventions on the *Felicity App*. Screening criteria was developed and quality appraisal was used to ensure only relevant articles were included in the review. However, more research should be done on whether mindfulness practices can improve academic performance and their impact on cognition.

## Data Availability

N/A

## DECLARATIONS

### Availability of data and material

N/A

### Code availability

N/A

### Authors’ contributions

JX devised and supervised the project and secured funding acquisition. AP and LN established the screening criteria and performed the searches. HJ and AL carried out the quality assessment and data extraction. AP, LN, HJ and AL equally contributed in data analysis and manuscript write up. JX and HJ refined and approved the manuscript.

## APPENDIX

## Appendix 1: Data Searches

| Database | Search | Results |
| --- | --- | --- |
| IEEE Xplore | (motivat*) OR (burnout) OR (student) OR (virtual) AND (mindfulness) | 65 |
| PsychInfo | (mindfulness) AND (motivation) OR (burnout) AND (student) AND (productivity) AND (intrinsic) AND (america) AND (virtual) AND (college) NOT (paid) | 664 |
| Web of Science | (motivation) OR (burnout) AND (student) AND (productivity) AND (intrinsic) AND (america) AND (virtual) AND (college) NOT (paid) | 19 |
| OVID | (mindfulness) OR (burnout) AND (student) AND (productivity) AND (intrinsic) AND (virtual) AND (college) | 96 |

## Appendix 2: Grey Literature Searches

| Database | Search | Results |
| --- | --- | --- |
| National Center for Biotechnology Information (NCBI) | (mindfulness) AND (motivation) OR (burnout) AND (student*) AND (productivity) AND (intrinsic) AND (america) AND (virtual) AND (college) NOT (paid) | 63 |
| Web of Science | (mindfulness) AND (motivation) OR (burnout) AND (student*) AND | 8 |
|  | (productivity) AND<br>(intrinsic) AND (america)<br>AND (virtual) AND (college)<br>NOT (paid) |  |

## Appendix 3: Study Characteristics

| First Author | Title | Country | Study design, duration, and participants | Type of Data | Outcome | Quality: Design/score |
| --- | --- | --- | --- | --- | --- | --- |
| Lin | Impact of mindfulness meditation intervention on academic performance | Taiwan | Quantitative non-randomized study; 2 hours for 3 months; first year undergraduate students in experimental group received mindfulness meditation training before training every week (n = 42) | Corresponding quiz, questionnaire to measure meditation depth and understand student satisfaction | Experimental results showed that MM significantly improves short-term academic performance (i.e. in-class quiz score) but does not significantly improve long-term academic performance (i.e. the SAs), questionnaire results reveal that most students enjoyed the MM process and agreed that MM intervention can help in-class learning. | 3/***** |
| Baird | Domain-specific enhancement of metacognitive ability following meditation training | USA | Quantitative randomized controlled trials; classes met for 45 min four times per week for 2 weeks; 50 undergraduate students were randomly assigned to either a meditation class (n = 26) or a nutrition class (n = 24) using a mixed factorial pretest-posttest design | Compliance with outside assignments assessed with daily journals, quantification of metacognitive ability | Compared with an active control group that elicited no change, we found that a 2-week meditation program significantly enhanced introspective accuracy, quantified by metacognitive judgments of cognition on a trial-by-trial basis, in a memory but not a perception domain. | 2/*** |
| Bamber | Mindfulness-based meditation to decrease stress and anxiety in college students: A narrative synthesis of the research | USA | Qualitative study; (exact time not specified); literature search reviewing research that tested effects of mindfulness meditation on stress and anxiety in college students (n = 57 studies) | Literature search | MBSR and MM interventions reduce stress & anxiety in college students; Self-reported stress decreased in majority of studies | 1/**** |
| Bamber | Effects of Mindfulness Meditation on College Student Anxiety: a Meta-Analysis. Mindfulness | USA | Qualitative study; (exact time not specified); Meta analysis examining mindfulness meditation on anxiety in college students (searched 11 electronic databases, hand searched select journals, and unpublished literature, located 25 primary studies, providing 28 comparisons, with 1492 participants) | Descriptive statistics comparing mindfulness-based interventions (MBIs) | MBIs have a large and significant effect in decreasing college students' anxiety. Despite researchers' emphasis on relationship/loving kindness and insightful meditations, these aspects of mindfulness meditation interventions did not influence MBI's effect on anxiety. | 1/**** |
| Banks | The protective effects of brief mindfulness meditation training | USA | Quantitative randomized controlled trials; 1 week at home mindfulness meditation intervention compared to an active control intervention; Eighty meditation-naïve undergraduate students from Nova Southeastern University participated for partial course credit and were randomly assigned to the MMT condition (n = 40) or RT condition (n = 40) | Automated Operation Span (AOSPAN), Task Unrelated Thoughts (TUTs), and Dundee Stress State Questionnaire (DSSQ) | Mindfulness meditation does not increase working memory or decrease mind wandering but does prevent stress related working memory impairments | 2/***** |
| Quek | Brief Mindfulness Breathing Exercises and Working Memory Capacity: Findings from Two Experimental Approaches | Singapore | Quantitative randomized controlled trials; single 15-min session; mindful attention session compared to mind-wandering exercise in a within-subjects experimental design (Study 1; N = 82) and a between-subjects experimental design (Study 2; N = 424) | Working memory capacity, span task | No difference between controls in working memory capacity tasks (operation span and symmetry span). Single session is not enough to enhance working memory capacity. | 2/**** |
| Rahl | Brief mindfulness meditation training reduces mind wandering: The critical role of acceptance | USA | Quantitative randomized controlled trial; incorporating instruction in both attention monitoring and acceptance, a mindfulness training condition incorporating attention monitoring instruction only, a relaxation training condition, or a reading control condition; 3-day brief mindfulness training condition; healthy young adults (n = 147) | 6-min Sustained Attention to Response Task (SART) | Mindfulness training and acceptance skills may facilitate emotion regulation on boring and frustrating sustained attention tasks that foster mind-wandering and may reduce mind wandering on the SART (sustained attention response task) | 2/***** |
| Shapiro | Cultivating mindfulness: effects on well-being | USA | Quantitative descriptive; college undergraduates in two groups - MBSR (n = 15), E.Easwaran's Eight Point Program (n = 14), or control (n = 15); pretest posttest and 8 week follow-up data were gathered on self-report outcome measures | Pretest, posttest, and 8-week follow-up data gathered on self-report outcome measures | Distinct meditation-based practices can increase mindfulness as measured by the MAAS and reduce perceived stress in participants. Single session is not enough to enhance working memory capacity. | 4/**** |
| Dvořáková | Promoting healthy transition to college through | USA | Quantitative randomized controlled trial; using Learning to BREATHE (L2B) program; 8 sessions over 6 weeks; freshmen | Mindful Attention Awareness Scale (MAAS) | Attending the workshops lead to significant improvements in students' life satisfaction, depression, anxiety, sleep issues and alcohol consequences. | 2/*** |
|  | mindfulness training with first-year college students: Pilot randomized controlled trial |  | recruited from residential halls (n = 109) |  |  |  |
| Jain | A randomized controlled trial of mindfulness meditation versus relaxation training: effects on distress, positive states of mind, rumination, and distraction | USA | Quantitative randomized controlled trial; effects of a 1-month mindfulness meditation versus somatic relaxation training; Full-time medical students, graduate nursing students, and undergraduate students majoring in pre medical or pre health studies (n = 81, 27 meditation, 24 relaxation, and 30 control; 15 men and 66 women) | Psychological distress, positive states of mind, distractive and ruminative thoughts and behaviors, and spiritual experience measured | Meditation and relaxation groups had decreases in distress and increases in positive moods over time compared to the control group. | 2/**** |
| Zeidan | Mindfulness meditation improves cognition: Evidence of brief mental training. Consciousness and Cognition | USA | Quantitative randomized controlled trial; four sessions of either meditation training or listening to a recorded book (n = 63) | Self report measures, cognitive measures | Both interventions were effective in mood improvement; only brief meditation training reduced fatigue, anxiety and increased mindfulness. 4 days of meditation training can enhance ability to sustain attention | 2/**** |
| Bellinger | Mindfulness, anxiety, and high-stakes mathematics performance in the laboratory and classroom | USA | Quantitative descriptive study; comparing performance in high pressure setting specifically completing modular arithmetic; freshman undergraduate engineering students enrolled in a calculus course (n = 112) | Marks from calculus course, mindfulness questionnaires (MMAS) | Mindfulness indirectly benefited math performance (reduced experience of state anxiety) and indirectly benefits students performance on high stakes quizzes and exams ( reduced test anxiety). Didn't impact performance on lower stakes assignments. | 4/**** |
| Morrison | Taming a wandering attention: short-form mindfulness training in student cohorts | USA | Quantitative randomized controlled trial; mindfulness training exercises; 7 hours over 7 weeks, University of Miami students (n = 58) | Performance on the sustained attention to response task (SART) and two working memory tasks (operation span, delayed recognition with distractors) | Mindfulness training (MT) participants had higher task accurate and self reported being more "on task" after 7 week training. MT didn't significantly benefit operation span task accuracy and doesn't improve working memory. | 2/**** |
| O'Driscoll | The effects of mindfulness-based interventions for health and social care undergraduate students - a systematic review of the literature | USA | Quantitative randomized controlled trial; systematic review to identify effects of mindfulness-based interventions to health and social care undergraduate studies; studies with medicine, nursing and psychology students that met inclusion criteria (n = 11) | Five Facet Mindfulness Questionnaire and the General Health Questionnaire | Benefits reported in participants' stress, mood and mindfulness levels, anxiety, well-being, self compassion and coping abilities. Further research with long-term follow-up is required to definitively conclude that mindfulness is an appropriate intervention to mentally prepare health and social care undergraduate students for their future careers. | 2/*** |
| Oman | Meditation lowers stress and supports forgiveness among college students: a randomized controlled trial | USA | Quantitative randomized controlled trial; two 8 week 90 min per week training program; college undergraduates in mindfulness-based stress reduction group n = 15), Easwaran's Eight-Point Program n = 14), or wait-list control (n = 15). | Self report outcome measures | No treatment differences between MBSR and EPP. Treated patients had significant benefits for stress and forgiveness. Evidence shows that meditation-based stress management practises reduce stress + enhance forgiveness among college students | 2/**** |
| Shapiro | Effects of mindfulness-based stress reduction on medical and premedical students. | USA | Quantitative randomized controlled trial; 7 week mindfulness-based intervention or wait list control group; 20 Fostering and Achieving Cultural Equity and Sensitivity students, 50 honors premedical students and 130 first and second-year medical students (n = 78) | Empathy Construct Rating Scale (ECRS), Hopkins Symptom Checklist 90, State-Trait Anxiety Inventory, Index of Core Spiritual Experiences | After MBSR interventions, the treated group had less depression, less state anxiety, less trait anxiety, a decrease in GSI, and increases in empathy and spirituality. | 2/**** |
| Brown | Mindfulness Enhances Episodic Memory Performance: Evidence from a Multimethod Investigation. | USA | Mixed media study 1 (n = 143) self reported state of mindful attention; study 2 (n = 93) brief training in focused attention for of mindfulness; study 3 (n = 57) mindfulness training generalized to free recall memory performance | Study 1: Mindful Attention Awareness Scale (MAAS) and Act with Awareness Subscale of the Five Factor Mindfulness Questionnaire; Study 2: 7-item Profile of Mood States | FFMQ results increased significantly between pre course and post course. Improvement was mainly seen in being less judgmental and less reactive towards thoughts, feelings, and emotions of others and particularly of oneself. | 5/***** |
| de Bruin | Mindfulness in higher education: awareness and attention in | Netherlands | Quantitative descriptive; assessed the effects of a mindfulness course in the curriculum of international students (n = 104) from 16 different countries at the | Five Facet Mindfulness Questionnaire (FFMQ) | The effect of breathing exercises was greater on the high math anxiety group. Effect of L-theanine showed greater calmness on the high math anxiety group but with low significance | 4/**** |
|  | university students increase during and after participation in a mindfulness curriculum course |  | University of Amsterdam; 7 weekly lectures |  |  |  |
| Brunyé | Learning to relax: Evaluating four brief interventions for overcoming the negative emotions accompanying math anxiety | USA | Quantitative randomized controlled trial; results of arithmetic task after practicing one of three short-term breathing exercises promoting focused attention, unfocused attention, or worry; Tufts University undergraduates with low vs. high math anxiety (n = 36) | Mathematics Anxiety Rating Scale (MARS), Mindful Attention Awareness Scale (MAAS), Five Facet Mindfulness Questionnaire (FFMQ) | Students assigned to practice mindfulness performed better on the first two quizzes than the control group. When both class sections had mindfulness meditation before Quiz 3, there was no significant difference between groups. Small doses of mindfulness training can briefly enhance students' knowledge retention of lecture content. | 2/***** |
| Calma-Birling | Does A Brief Mindfulness Intervention Impact Quiz Performance? Psychology Learning and Teaching | USA | Quantitative randomized controlled trial; effects of a brief mindfulness intervention on quiz performance; two sections of an undergraduate upper-level human development class (n = 67); met every Tuesday and Thursday for an hour and twenty mins | Three quizzes, three exams, state mindfulness scale | Mood ratings and led to better quiz scores but not better final exam scores. The interventions did not result in better same-day learning, but students performed better on the final exam items from intervention days, particularly the cognitive reappraisal day. | 2/***** |
| Cavanagh | A multicourse, multi semester investigation of the impact of cognitive reappraisal and mindfulness instruction on short- and long-term learning in the college classroom | USA | Quantitative randomized controlled trial; 3 intervention days that participants rated their emotions; course taught over Spring 2017 and Fall 2017 semesters at a small liberal arts Catholic college in New England (n = 226) | Mood ratings on 12 measurement occasions, self reported perceptions of study, quiz and final exam scores | The interventions did not result in better same-day learning, but students performed better on the final exam items from intervention days, particularly the cognitive reappraisal day. | 2/***** |
| Hoffmann Gurka | Mindfulness meditation for college students: A study of its utility and promotion of its practice post treatment | USA | Quantitative randomized controlled trial; mindfulness-based stress reduction program for 4 weeks that taught two forms of mindfulness-based stress management techniques; college students from major university (n = 123) | Test scores | Mindfulness training seemed to help student interpreters become more aware of and regulate their own emotions and attention, experience greater equanimity and less reactivity, and become kinder toward themselves, including when they were interpreting. Some participants, however, may have been over-attributing to mindfulness the progress they felt they were making in their interpreting classes. | 2/***** |
| Johnson | Effect of Mindfulness Training on Interpretation Exam Performance in Graduate Students in Interpreting | USA | Mixed method design; effect of short-term mindfulness training on exam performance; 4 week 12 hour mindfulness training across 7 language programs (n = 67) | Pretests and posttests for consecutive interpreting exam performance, Cognitive and Affective Mindfulness Scale- Revised (CAMS-R), perceived stress (PSS-10), and attention (d2 Test of Attention) | Mindfulness training seemed to help student interpreters become more aware of and regulate their own emotions and attention, experience greater equanimity and less reactivity, and become kinder toward themselves, including when they were interpreting. | 5/*** |
| Metz | The Effectiveness of the Learning to BREATHE Program on Adolescent Emotion Regulation. Research in Human Development | USA | Quantitative non-randomized study; effectiveness of the mindfulness-based training program Learning 2 BREATHE on adolescent emotion regulation, perceived efficacy in affective regulation, perceived stress, and somatic symptoms; regular education high school students (n = 216) | Voluntary self-report survey, 13-item Psychosomatic Complaints Scale, 14-item Affective Self-Regulatory Efficacy Scale (ASRES) | Learning 2 BREATHE has a positive effect on measures of emotional regulation, self-regulation efficacy, psychosomatic complaints, and self-report stress level. | 3/***** |
| Potek | Mindfulness as a school-based prevention program and its effect on adolescent stress, anxiety and emotion regulation. | USA | Quantitative randomized controlled trial; explored effectiveness of Learning 2 BREATHE program; 30 students in two high schools (one rural and one urban) | Multidimensional Anxiety Scale for Children (MASC), Follow-up (F1) data after program implementation and Follow-up (F2) data after program completion | Significant reduction in self report rated symptoms of anxiety. No moderating effects of homework. No differential outcomes based on school site differences. | 2/***** |
| Bernardez | Effects of Mindfulness on Conceptual Modeling Performance: a Series of Experiments. IEEE Transactions on | USA | Quantitative non-randomized study; several weeks of mindfulness practice over three academic years from 2013 to 2016; second year students of Degree in Software Engineering at the University of Seville (n = 130) | State Trait Anxiety Inventory questionnaire, four exam scores | Subjects who practiced mindfulness developed slightly better conceptual models (their quality was 8.16% higher) and they did it faster (they were 46.67% more productive) than the control group. | 3/**** |
|  | Software Engineering |  |  |  |  |  |
| Sankoh | Mindfulness in medicine: Modified Mindfulness-Based Stress Reduction (MBSR) Program among Future Doctors | USA | Quantitative descriptive; participants assigned to one of three groups: 8 week mindfulness course (n = 16), 8 week general stress reduction course without specific mindfulness instruction (n = 10), or no treatment; premedical and medical student (n = 14) | Mindfulness Attention Awareness Scale (MAAS), the Yoni Test of cognitive and affective Theory of Mind (Yoni), Maslach Burnout Inventory – Student Survey (MBI-SS), Mayer-Salovey-Carusuo Emotional Intelligence Test (MSCEIT) | Participants in the mindfulness intervention expressed significantly less student burnout when compared to the active and inactive control group participants. But didn't produce significantly higher levels of mindfulness or emotional intelligence among its participants when compared to control participants. | 4/**** |
| Stein | Impact of a 3-minute mindfulness-based exercise on anxiety and academic performance | USA | Quantitative non-randomized study; 3 minute mindfulness exercise at beginning of each class for a period of 6 weeks; anxiety and academic performance assessed; first year college students (n = 105) | State-Trait Anxiety Inventory questionnaire, four exam scores | Students who participated in mindfulness exercise reported lower state anxiety levels (measured by State Trait Anxiety Inventory). Academic performance not significantly impacted by engaging in mindfulness exercise | 3/**** |
| Weis | Mindfulness as a way to cope with COVID-19-related stress and anxiety | USA | Quantitative non-randomized study; determine whether Koru Mindfulness-based group therapy could help students cope with COVID 19 related stress and anxiety; 4 week program in psychology research class at midwestern university; undergraduate students (n = 32) | The Cognitive and Affective Mindfulness Scale-Revised, Self-Compassion Scale, The Medical Outcomes Study Sleep Scale, The Perceived Stress Scale, The Conners Continuous Performance Test 3rd Edition | KM participants reported greater mindfulness and self-compassion and less stress, anxiety and sleep problems than controls. KM participants showed superior functioning on performance based measures of attention. Most gains were maintained over time. | 3/**** |
| Yamada | The Impact of Mindful Awareness Practices on College Student Health, Well-Being, and Capacity for | USA | Quantitative non-randomized study; utility of mindful awareness practices (MAPs) in terms of enhancing student learning in the college classroom, as well as improving psychological well-being; 10 minute MAP | Freiburg Mindfulness Inventory, Sense of Capacity for learning in the classroom | Mindfulness intervention did not lead to significant improvement in academic performance across the semester. But 81% of students self-reported positive effects of MAPs on their learning. | 3/***** |
|  | Learning: A Pilot Study. Psychology Learning and Teaching |  | at beginning of every class; undergraduate students (mindfulness group n = 37; control group n = 23) |  |  |  |
| Yamaji | Effects of Mindful Somatic Psychoeducation for Japanese College Students | Japan | Quantitative randomized controlled trial; role of somatic psychoeducation in developing somatic awareness and trust in the organism, and enhance integral functioning that included general mindfulness, stress resilience, interpersonal empathy, responsibility for self-care, and generic skill; sophomore students at Japanese university (n = 59) | Self report questionnaires, performance measures | Experimental course was successful in cultivating somatic awareness and the observing component of mindfulness, but not in other dependent variables (empathy, resilience, responsibility for self-care, trust in the organism, attitudinal components of mindfulness, and the self-evaluation of generic skills) | 2/**** |
| Young | The impact of a well-being program imbedded in university classes: the importance of valuing happiness, baseline well-being and practice frequency | Australia | Qualitative study; effect of positive psychology well-being intervention; 3 separate studies and over 3 years; undergraduate students in psychology class (n = 469) | Mental Health Continuum-Short Form (MHC-SF), Negative Affect Schedule (PANAS), Valuing Happiness Scale | Intervention outcomes across three years indicate improvements in well-being, positive affect, negative affect and clinical well-being categories. Intervention buffered against semester stress. Practice frequency had little effect on well-being, however well-being gains were higher for students with low baseline well-being and those who valued happiness more – suggesting positive education interventions have greater value for some students | 1/***** |

## Appendix 4: Excluded Studies

Excluded Studies from Quality Appraisal.

Appendix 4: Excluded Studies Excluded Studies from Quality Appraisal
| Author | Title of Article |
| --- | --- |
| Beatriz Bernardez, Amador Duran Toro, Jose Antonio Prejo, Antonio Ruiz-Cortes | An experimental replication on the effect of the practice of mindfulness in conceptual modeling performance |
| Karen Bluth, Rebecca A. Campo, Sarah Pruteanu-Malinici, Amanda Reams, Michael Mullarkey, and Patricia C. Broderick | A School-Based Mindfulness Pilot Study for Ethnically Diverse At-Risk Adolescents |
| Joanna J Arch, Michelle G Craske | Mechanisms of mindfulness: Emotion regulation following a focused breathing induction. Behaviour Research and Therapy |
| Nicole Wolff, Christian Best | Short-term Smartphone App-Based Focused Attention Meditation Diminishes Cognitive Flexibility |
| Kamila Dvorakova | Using Contemplative Practices to Promote College Students' Socioemotional Competencies and Healthy Transition to College: A Conceptual and Empirical Analysis |

Excluded Studies from Hand Searches.

Excluded Studies from Hand Searches
| Author | Title of Article |
| --- | --- |
| Destany Calma-Birling, Regan A. R. Gurung | Does A Brief Mindfulness Intervention Impact Quiz Performance? |
| Brockman R, Ciarrochi J, Parker P, Kashdan T. | Emotion regulation strategies in daily life: mindfulness, cognitive reappraisal and emotion suppression. |
| Ida H Bennike, Anders Wieghorst, Ulrich Kirk | Online-based Mindfulness Training Reduces Behavioral Markers of Mind Wandering |
| Chan RW, Lushington K, Immink MA | States of focused attention and sequential action: A comparison of single session meditation and computerised attention task influences on top-down control during sequence learning. |
| Felicia Huppert Daniel Johnson | A controlled trial of mindfulness training in schools: The importance of practice for an impact on well-being |
| Allen, M., Dietz, M., Blair, K. S., van Beek, M., Rees, G., Vestergaard-Poulsen, P., Lutz, A., Roepstorff, A. | Cognitive-affective neural plasticity following active-controlled mindfulness |
| Britt, M. E. | Effect of a mindfulness intervention on community college students' writing apprehension and writing performance. |
| Broderick P. | Learning to BREATHE: A mindfulness curriculum for adolescents to cultivate emotion regulation, attention, and performance. |
| Burke, C. | Mindfulness-based approaches with children and adolescents: a preliminary review of current research in an emergent field. |
| Chiesa A., Calati R., & Serretti A. | Does mindfulness training improve cognitive abilities? A systematic review of neuropsychological findings. |
| Pogrebtsova E, Craig J, Chris A, O'Shea D, González-Morales MG. | Exploring daily affective changes in university students with a mindful positive reappraisal intervention: A daily diary randomized controlled trial. |
| Franco, C., Manas, I., Cangas, A., & | The application of mindfulness with students of secondary school: |
| Gallego, J. | Results on the academic performance, self-concept and anxiety |
| Galante, J., Dufour, G., Vainre, M., Wagner, A., Stochl, J., Benton, A., & Jones, P. | A mindfulness-based intervention to increase resilience to stress in university students. |
| Galante, J., Stochl, J., Dufour, G., Vainre, M., Wagner, A., & Jones, P. | Effectiveness of providing university students with a mindfulness-based intervention to increase resilience to stress. |
| Greeson, J., Juberg, M., Maytan, M., James, K., & Rogers, H. | A randomized controlled trial of Koru. |
| Hindman, R., Glass, C., Arnkoff, D., & Maron, D. | A comparison of formal and informal mindfulness programs for stress reduction in university students. |
| Holland, D. | Contemplative education in unexpected places: Teaching mindfulness in Arkansas and Austria. |
| J. T. Ramsburg and R. J. Youmans | Meditation in the higher– education classroom: Meditation training improves student knowledge retention during lectures |
| Jha, A., Krompinger, J., & Baime, M. | Mindfulness training modifies subsystems of attention. |
| Kang, Y., Choi, S. & Ryu, E. | The effectiveness of a stress coping program based on mindfulness meditation on the stress, anxiety, and depression experienced by nursing students in Korea. |
| Broderick P. C. et al. | Learning to BREATHE: A pilot trial of a mindfulness curriculum for adolescents. |
| M. D. Mrazek, M. S. Franklin, D. T. Phillips, B. Baird, and J. W. Schooler, | Mindfulness training improves working memory capacity and GRE performance while reducing mind wandering |
| Metz S, Frank J, Reibel D, Cantrell T, Sanders R, Broderick P. | The effectiveness of the Learning to BREATHE program on adolescent emotion regulation. Research in Human Development. |
| Michael de Vibe, Ida Solhaug, et. al | Mindfulness training for stress management: a randomised controlled study of medical and psychology student |
| Mrazek MD, Franklin MS, Phillips DT, Baird B, Schooler JW. | Mindfulness training improves working memory capacity and GRE performance while reducing mind wandering. |
| Moore A., Gruber T., Deroose J., Malinowski P. | Regular, brief mindfulness meditation practice improves electrophysiological markers of attentional control. |
| N. Barnes, P. Hattan, D. S. Black, and Z. Schuman-Olivier | An examination of mindfulness–based programs in us medical schools |
| Ramsburg J. T., Youmans R. J. | Meditation in the higher-education classroom: meditation training improves student knowledge retention during lectures. |
| Regehr, C., Glancy, D., & Pitts, A. | Interventions to reduce stress in university students: A review and meta-analysis. |
| Rosenzweig, S., Reibel, D. K., Greeson, J. M., Brainard, G. C., & Hojat, M. | Mindfulnessbased stress reduction lowers psychological distress in medical students. |
| Sauer-Zavala, S. E., Walsh, E. C., Eisenlohr-Moul, T. A., & Lykins, E. L. B. | Comparing mindfulness-based intervention strategies: Differential effects of sitting meditation, body scan, and mindful yoga. |
| Valentine, E. R., & Sweet, P. L. G. | Meditation and attention: A comparison of the effects of concentrative and mindfulness meditation on sustained attention. |
| Vettese, L. C., Toneatto, T., Stea, J. N., Nguyen, L., & Wang, J. J. | Do mindfulness meditation participants do their homework? And does it make a difference? A review of the empirical evidence. |
| Warnecke, E., Quinn, S., Ogden, K., Towle, N., & Nelson, M. R. | A randomised controlled trial of the effects of mindfulness practice on medical student stress levels |
| Yusufov M, Nicoloro-SantaBarbara J, Grey NE, Moyer A, & Lobel M | Meta-analytic evaluation of stress reduction interventions for undergraduate and graduate students. |
| Zenner, C. , S.Herrleben-Kurz, and H.Walach | Mindfulness-based Interventions in Schools – a Systematic Review and Meta-Analysis. |

Excluded Studies from Full Text.

Excluded Studies from Full Text
| Author | Title of Article |
| --- | --- |
| A. A. Alvarez | Urban student perspectives on classroom-based daily mindfulness practices |
| A. J. Amutio-Kareaga, Clemente Franco Gazquez Linares, Jose Jesus Manas, Israel Manas | Learning mindfulness for enhancing relaxation and self- efficacy on academic performance in high school students |
| A. Y. S. Aranega, R. C. Perez, C. G. | Mindfulness' effects on undergraduates' perception of self-knowledge and stress levels |
| P. P. M. P.-M. J. Avery | Special Session - Lazy Wisdom: Teaching Methods and Realization Activities Enabling Students to Acquire Reawakened Receptive Modes of Knowledge |
| B. Ü. M. B. T. H. Ç. D. Ayberkin | Relations of attention and meditation level with learning in engineering education |
| S. R. Babbar, K. Williams, K. | Addressing Obstetrics and Gynecology Trainee Burnout Using a Yoga-Based Wellness Initiative During Dedicated Education Time |
| M. D. S. Bamber, Joanne Kraenzle | College students' perceptions of mindfulness-based interventions: A |
|  | narrative review of the qualitative research |
| M. V. Beardsley, M. Portero-Tresserra, M. Hernandez-Leo, D. | ClassMood App: A Classroom Orchestration Tool for Identifying and Influencing Student Moods |
| M. M. K. Beattie, H. M. Volanen, S. M. Knittle, K. P. Hankonen, N. E. | Social Cognitions and Mental Health as Predictors of Adolescents' Mindfulness Practice |
| J. Bhatti | High Test Anxiety in Chiropractic Students: Assessment of an Educational Intervention |
| L. Birnbaum | The use of mindfulness training to create an 'accompanying place' for social work students |
| P. C. M. Broderick, Stacie | Learning to BREATHE: A pilot trial of a mindfulness curriculum for adolescents |
| P. C. M. Broderick, Stacie M. | Working on the inside: Mindfulness for adolescents |
| Y. L. E. N. A. R. T. D. A. S. S. T. N. D. J. D. I. Chamindi | Smartphone-based Approach to Enhance Mindfulness Among Undergraduates with Stress |
| S. C. Chase-Cantarini, Glenda | Introducing mindfulness moments in the classroom |
| C.-C. Chen | An implementation of therapeutic-based art pedagogy: Enhancing culturally diverse students' self-esteem |
| S. R. Clark | The impact of self-regulated attention control on the amount of time spent in flow |
| L. G. Corti, C. | Mindfulness and Coaching to Improve Learning Abilities in University Students: A Pilot Study |
| J. P. Croskey, II | Undergraduates' experiences with mindfulness practice: A qualitative study |
| C. P. Davenport, Francesco | Mindful learning: A case study of Langerian mindfulness in schools |
| S. N. P. M. V. Desai | A Survey on Effects of Various Meditation Interventions on Overall Performance of College Students |
| P. L. H. Dobkin, Tom A. | Teaching mindfulness in medical school: Where are we now and where are we going? |
| L. B. Dye, Monica Galloway Wolf, Cheryl | Teaching mindfulness for the self-care and well-being of counselors-in-training |
| L. Edwards | Combining biofeedback and mindfulness in education |
| B. E. Elphinstone, P. Whitehead, R. | Greater autonomous motivation for study and basic psychological need |
|  | satisfaction by being presently aware and 'letting go': An exploration of mindful attention and nonattachment |
| B. W. Elphinstone, R. Bates, G. | 'Letting go' and flourishing in study: An investigation of the indirect relationship between nonattachment and grades via psychological well-being |
| E. H. W. Eustis, Sarah Krill Morgan, Lucas P. Graham, Jessica R. Hayes-Skelton, Sarah A. Roemer, Lizabeth | Development, acceptability, and effectiveness of an acceptance-based behavioral stress/anxiety management workshop for university students |
| A. L. T. Eva, Natalie M. | Learning to BREATHE: A pilot study of a mindfulness-based intervention to support marginalized youth |
| N. L. L. Fonger, Kien | The promise of mindfulness as a proposed intervention to alleviate the delimiting effects of math anxiety |
| L. G. Forbes, D. Johnson, S. K. | Investigating Adherence to an Online Introductory Mindfulness Program |
| M. Fowler | Mindful discipline for distressed learners |
| C. S. Franco, E. Amutio, A. Manas, I. | Improving motivation in Latin American immigrants through a mindfulness-based program: a randomized study |
| J. M. Froiland | Promoting gratitude and positive feelings about learning among young adults |
| N. T. Gabana | Exploring the effects of a gratitude intervention with college student-athletes |
| B. M. E. Galla, M. V. Fiore, H. M. | Mindfulness predicts academic diligence in the face of boredom |
| B. A. Gaudiano | Mindfulness: Nonclinical applications of mindfulness: Adaptations for school, work, sports, health, and general well-being. Vol. IV |
| A. B. Gockel, David James, Susan Bryer, Ellen | Introducing mindfulness as a self-care and clinical training strategy for beginning social work students |
| A. C. Gockel, Theresa Malove, Shirley James, Susan | Mindfulness as clinical training: Student perspectives on the utility of mindfulness training in fostering clinical intervention skills |
| E. C. Goh | Integrating mindfulness and reflection in the teaching and learning of listening skills for undergraduate social work students in Singapore |
| F. R. K. Goodman, Todd B. | Behind the scenes of clinical research: Lessons from a mindfulness intervention with student-athletes |
| S. L. Gu, Yawen Liang, Fei Feng, Rou | The mediating effects of coping style on the effects of breath count |
| Zeng, Zhi Wang, Fushun | mindfulness training on depressive symptoms among international students in China |
| W. B. S. Hansen, L. M. | Specialized Smartphone Intervention Apps: Review of 2014 to 2018 NIH Funded Grants |
| M. Hilburn-Arnold | Turning to creativity: A grounded theory approach towards understanding the relationship between wellness and the arts for adolescents |
| A. B. Hjeltne, Per-Einar Moltu, Christian Dundas, Ingrid | Facing the fear of failure: An explorative qualitative study of client experiences in a mindfulness-based stress reduction program for university students with academic evaluation anxiety |
| A. J. B. Howell, Karen | Relations among mindfulness, achievement-related self-regulation, and achievement emotions |
| A. L. Huang | Mindfulness support group for college students: Combatting their fears and stresses |
| E. T. Hulme, Christy | Mindfulness in student affairs practice |
| T. A. Ivey | Perceived effectiveness and application of mindfulness practices in education: A qualitative study |
| P. A. D. Jennings, Anthony A. Mischenko, Polina P. | Where are we now? Where are we going?: Preparing our students for an uncertain future |
| S. J. V. Kass, Lisa A. Mikulas, William L. Legan, Shauna Bumgarner, David | Effects of mindfulness training on simulated driving: Preliminary results |
| D. C. Kerrigan, Victoria King, Melissa Holman, Emily Joffe, Alain Sibinga, Erica | There is no performance, there is just this moment: The role of mindfulness instruction in promoting health and well-being among students at a highly-ranked university in the United States |
| E. G. Kimble | Acceptability and outcomes of a guided-meditation intervention for school-age second language learners |
| H. R. Kindel | Toward expert clinicians: The effects of teaching mindfulness in physical therapy education |
| S. King | A case study of a yoga and meditation intervention in an urban school: A complex web of relationships and resilience in the search for student well-being |
| K. M. L. Kraemer, Christina M. O'Bryan, Emily M. Mysinger, Erica Cotton, Sian | Mind-body skills training to improve distress tolerance in medical students: A pilot study |
| C. P. B. Kreutzer, Clint A. | Making games for health engaging: The influence of cognitive skills |
| E. J. Langer | Mindfulness forward and back |
| D. Laureiro-Martinez | Cognitive control capabilities, routinization propensity, and decision-making performance |
| S. Lauricella | Mindfulness meditation with undergraduates in face-to-face and digital practice: A formative analysis |
| N. M. Lemon, Sharon | Mindfulness in the academy: Practices and perspectives from scholars |
| E. L. B. Lykins | Effects of mindfulness and meditation experience on cognitive and emotional functioning and ego depletion |
| M. G. McAlister | Deepening awareness: The integration of mindfulness practices in United States high schools |
| C. J. E. Miller, Katelyn Scavone, Antonette | The feasibility of bringing brief mindfulness-based training to the university classroom |
| V. D. Mirisse | Education for mindfulness: From the diary of a monk |
| E. R. V. K. Mittal | Effect of different music genre: Attention vs. meditation |
| M. D. Z. Mrazek, Claire M. Gross, Madeleine E. Mrazek, Alissa J. Phillips, Dawa T. Schooler, Jonathan W. | Mindfulness in education: Enhancing academic achievement and student well-being by reducing mind-wandering |
| M. B. Napoli, Robin | From theory toward empathic self-care: Creating a mindful classroom for social work students |
| G. M. Nixon, D. Craig, L. Nevejan, A. Regan-Addis, H. | Studies in mindfulness: widening the field for all involved in pastoral care |
| M. T. D. R. Nunez, F. J. C. Sanchez, R. C. Aranega, A. Y. | Developing management skills through experiential learning: the effectiveness of outdoor training and mindfulness |
| D. A. O'Brien | Using mindfulness meditation intermixed with humor to reduce anxiety among nursing students during clinical practice |
| L. C. C. F. F. B. R. C. Y. F. V. V. S. A. B. A. R. Palaniappan | Effect of Mindfulness Meditation toward Improvement of Concentration based on Heart Rate Variability |
| K. A. Parish | Quieting the cacophony of the mind: The role of mindfulness in adult learning |
| A. R. Rao | A novel STEAM approach: Using cinematic meditation exercises to motivate students and predict performance in an engineering class |
| D. T. Reid | Teaching mindfulness to occupational therapy students: Pilot evaluation of an online curriculum |
| S. Rongmuang | A mixed-methods study: The effect of embodied learning on nursing students' presence, well-being, relationships with patients, and learning experience |
| C. J. R.-C. Rybak, Lori A. | The teaching well: Experience, education and counselling |
| J. M. Sampl, T. Furtner, M. R. | A Randomized Controlled Pilot Intervention Study of a Mindfulness-Based Self-Leadership Training (MBSLT) on Stress and Performance |
| S. V. G. Sapthiang, William Shonin, Edo | Mindfulness in schools: A health promotion approach to improving adolescent mental health |
| D. L. O. Schussler, Yoonkyung Mahfouz, Julia Levitan, Joseph Frank, Jennifer L. Broderick, Patricia C. Mitra, Joy L. Berrena, Elaine Kohler, Kimberly Greenberg, Mark T. | Stress and well-being: A systematic case study of adolescents' experiences in a mindfulness-based program |
| S. H. Shannon, D. Haughey, T. Leavey, G. McGeown, C. Breslin, G. | Effects of a Mental Health Intervention in Athletes: Applying Self-Determination Theory |
| I. E. Solhaug, Thor E. de Vibe, Michael Haavind, Hanne Friborg, Oddgeir Sorlie, Tore Rosenvinge, Jan H. | Medical and psychology student's experiences in learning mindfulness: Benefits, paradoxes, and pitfalls |
| B. K. Soulakova, Alexandr Butzer, Bethany Winkler, Petr | Meta-review on the effectiveness of classroom-based psychological interventions aimed at improving student mental health and well-being, and preventing mental illness |
| R. I. Teper, M. | Meditation, mindfulness and executive control: the importance of emotional acceptance and brain-based performance monitoring |
| I. M. Thomas | The experience of mindfulness and learning: A qualitative research study |
| M. J. Trotter | Effects of participation in a Mindfulness-based Stress Reduction program on college students' Psychological Well-Being |
| Y. V. Z. M. J. Venkatesh | Democratizing Engineering Education Through Contemplative and Mindfulness Practices |
| V. R. Viciano, A. B. F. Linares, M. Espejo, T. Puertas, P. Chacon, R. | The University Studies and the Mindfulness. A Systematic Review |
| M. T. Vogel Trautt | Exploring the malleability of executive function through a mindful lens |
| X. G. Wang, Liuna Zhou, Kexin Ye, Lijuan Ma, Yinglin Zhang, Shuhao | Mindful learning can promote connectedness to nature: Implicit and explicit evidence |
| L. B. Waters, Adam Ridd, Amanda Allen, Kelly | Contemplative education: A systematic, evidence-based review of the effect of meditation interventions in schools |
| C.-Q. S. Zhang, Gangyan Duan, Yanping Lyu, Yaojun Keatley, David A. Chan, Derwin K. | The effects of mindfulness training on beginners' skill acquisition in dart throwing: A randomized controlled trial |
| Fischer, EV; Hebbeler, J | Modes. A Multi-sensory Media Experience for Stress Reduction |
| Sílvia Lopes, Armanda Pereira, Paula Magalhães, André Oliveira, and Pedro Rosário | *Gamification*: focus on the strategies being implemented in interventions: a systematic review protocol |
| Manuel Schmidt-Kraepelin, Philipp A Toussaint, Scott Thiebes, Juho Hamari, Ali Sunyaev, | *Archetypes of Gamification: Analysis of mHealth Apps* |

